# Evaluating the causal relationship between educational attainment and mental health

**DOI:** 10.1101/2023.01.26.23285029

**Authors:** Perline A. Demange, Dorret I. Boomsma, Elsje van Bergen, Michel G. Nivard

## Abstract

We investigate the causal relationship between educational attainment (EA) and mental health using two research designs. First, we compare the relationship between EA and 18 psychiatric diagnoses within sibship in Dutch national registry data (N=1.7 million), thereby controlling for unmeasured familial factors. Second, we apply two-sample Mendelian Randomization, which uses genetic variants related to EA or psychiatric diagnosis as instrumental variables, to test whether there is a causal relation in either direction. Our results suggest that lower levels of EA causally increase the risk of MDD, ADHD, alcohol dependence, GAD and PTSD diagnoses. We also find evidence of a causal effect of ADHD on EA. For schizophrenia, anorexia nervosa, OCD, and bipolar disorder, results were inconsistent across the different approaches, highlighting the importance of using multiple research designs to understand complex relationships such as between EA and mental health.

## Introduction

Over 17% of the population of the European Union is diagnosed with a mental disorder (2016 estimates, OECD, 2018). Psychiatric diagnoses are estimated to account for 25% of years lived with disability worldwide^2^. The risk of being diagnosed with a mental disorder is higher among those with lower educational attainment (EA)^3–5^. If the relationship between education and mental diagnoses is indeed causal, educational policies primarily aimed at improving educational outcomes could also lead to improved mental health.

Most prior studies of the relationship between education and mental health are correlational. This correlation could therefore reflect confounding factors influencing both education and mental health. While known and perfectly measured confounders can be controlled, unknown and unmeasured confounders cannot. Additionally, the correlation between EA and mental health might also be explained by reverse causation, as early onset of psychiatric symptoms may hamper subsequent school attendance and performance^6^. Randomized experiments in which education is altered would avoid bias due to confounding or reverse causation, but experiments at the required scale are not feasible for practical and ethical reasons.

As an alternative, we rely on two natural experiments that account for confounding and/or reverse causation in different manners: within-sibship regression and mendelian randomization (MR).

In the Dutch population registry (N=1.7 million siblings born between 1965 and 1985), we test whether differences in EA between siblings relate to differences in their risk of psychiatric diagnoses between 2011 and 2016. A core assumption of the within-sibship design is that siblings constitute a well-matched case-control group^7,8^. Siblings are comparable for many factors that might play a role in both EA and mental health, for example the family, school, and neighbourhood environment, and 50% of their segregating genome. By comparing siblings, we obtain estimates of the association of education with psychiatric diagnoses and care expenditures, controlled for unmeasured confounders shared by the siblings. While within-sibship estimates can increase or decrease our confidence in the presence of a causal relation, they are insufficient. Confounders not shared between siblings such as differential experiences, the other 50% of the segregating genome, but also non-shared measurement error might bias the estimate^9^. Within-sibship association does not offer evidence of a causal direction, and while timing of events may suggest direction, unmeasured prodromal signs and experiences anterior to graduation might affect the association.

To mitigate the uncertainties introduced by limitations of the within-sibship design, we also apply mendelian randomization (MR). In MR, genetic variants that are robustly associated with the exposure are used as instrumental variables. MR’s core assumptions are: (1) some variants are associated with the exposure (EA), (2) these variants are related to mental health only via their effect on educational success, and (3) these variants do not correlate with any confounders of the relationship between education and mental health^10^. When assumptions are met, MR estimates the causal effect of education on mental health, even in presence of confounding and measurement error^11^. Reverse causation can be empirically evaluated by running two sets of MR analyses: one with variants related to EA as exposure and variants related to psychiatric diagnoses as outcome, and the reverse analysis. We apply two-sample MR^12^, which uses genetic effect estimates from existing well-powered genome-wide association studies (GWASs) of EA^13^ and of psychiatric diagnoses in European-ancestry samples. To minimize the influence of pleiotropy (i.e. one genetic variant affects many traits), we used additional weak-instrument- and pleiotropy-robust MR methods^14–16^. To partly mitigate the influence of assortative mating, population stratification, and gene-environment correlation we performed MR based on genetic associations with EA obtained in a within-sibship GWAS^17^. Mendelian randomization applied to discrete or (ordered) categorical traits, such as EA and psychiatric diagnoses, has an additional notable caveat: interpretation^18^. If we assume that genetic variants influence categorical variables via their effects on the underlying liability, MR estimates the effect of the liability for higher EA on the liability for being diagnosed, while the within-sibship design estimates the effect of the observed exposure on the risk of the observed outcome.

Only a few prior quasi-causal experiments investigate EA and psychiatric diagnoses, and obtained mixed results. Studies that compared monozygotic twin pairs with discordant educational outcomes found evidence consistent with a causal association between EA and depressive symptoms^19^, while others did not^20,21^. Similarly, MR studies showed mixed evidence for a negative effect of EA on the risk for depression diagnoses^22–24^ but no reverse effect^25^. MR studies suggested a negative effect of EA on ADHD, no effect on PTSD and schizophrenia, but also a positive effect on schizophrenia, bipolar disorder, anorexia, autism and anxiety^26–28^. Within-sibship studies of psychiatric diagnoses and EA are rare, and even rarer in population registries^29,30^.

The reliance on quasi-causal methods, though preferred over observational association, is still imperfect, and calls for epistemic humility when interpreting or generalizing putative causal association. We reduce uncertainty on the nature of the relation between education and mental health by triangulating across two preregistered quasi-causal methods relying on different underlying assumptions, whose violation would give rise to different biases^31^. Consistent findings strengthen our confidence. Inconsistent findings raise scepticism yet are also valuable: we interpret these differences in the light of the different assumptions of each method to help us hypothesize on the mechanisms in the relation between education and mental diagnoses.

## Results

In the following, we focus on diagnoses for which large-scale GWASs are available. Results for all diagnoses described in **Table 1** are available in **Supplementary Note and Tables**.

**Table 1.** Prevalence of diagnoses in the Dutch population registry (CBS) between 2011 and 2016 and source of GWAS for matching diagnoses. The frequency of DSM-IV Diagnoses in the Dutch population registry registered in second-line/specialized mental health care, both as primary or secondary diagnoses. The definition of the disorder in the equivalent GWAS does not always perfectly align with the diagnosis in CBS, see **Supplementary Table 20** for details of GWASs used. Subsamples are relevant subsets of the Dutch population registry that met criteria for inclusion in the study. Inclusion is further described in the method section and the chart **Supplementary Figure 1**. EA: educational attainment. SD: Standard deviation.

|  |  | Subsamples of individuals in CBS born between 1965 and 1985 |  |  |  |  |
| --- | --- | --- | --- | --- | --- | --- |
|  |  | All (1) | All with EA (2) | Siblings (3) | Siblings with EA (4) |  |
| Total sample size |  | 6,539,767 | 3,305,733 | 3,234,923 | 1,743,032 |  |
| EA average (SD) |  | – | 14.7 (3.4) | – | 15.4 (2.8) |  |
| Percentage of the population diagnosed between 2011 and 2016 | Individuals without any disorder | 93.86 | 88.82 | 91.02 | 89.17 |  |
|  | Individuals with at least one disorder | 7.14 | 11.18 | 8.98 | 10.83 | GWAS |
|  | Attention deficit hyperactivity disorder (ADHD) | 0.93 | 1.53 | 1.31 | 1.62 | Demontis |
|  | Autism spectrum disorder (ASD) | 0.35 | 0.55 | 0.51 | 0.61 | Grove |
|  | Alcohol dependence | 0.92 | 1.41 | 1.12 | 1.30 | Walters |
|  | Schizophrenia/schizophreniform/schizoaffective disorders | 0.39 | 0.52 | 0.48 | 0.51 | – |
|  | Schizophrenia | 0.31 | 0.42 | 0.38 | 0.41 | Trubetskoy |
|  | Major depressive disorder (MDD) | 2.81 | 4.42 | 3.34 | 4.08 | Howard |
|  | Bipolar disorder | 0.22 | 0.33 | 0.29 | 0.34 | Mullins |
|  | Bipolar I | 0.15 | 0.22 | 0.19 | 0.23 | Mullins |
|  | Bipolar II | 0.08 | 0.13 | 0.11 | 0.14 | Mullins |
|  | Anxiety disorders | 2.50 | 3.94 | 3.01 | 3.70 |  |
|  | Generalized anxiety (GAD) | 0.42 | 0.67 | 0.56 | 0.70 | Purves |
|  | Panic | 0.64 | 1.00 | 0.81 | 0.97 | – |
|  | Phobia | 0.69 | 1.10 | 0.91 | 1.12 | – |
|  | Obsessive-compulsive (OCD) | 0.28 | 0.42 | 0.38 | 0.45 | Arnold |
|  | Post-traumatic stress (PTSD) | 1.16 | 1.84 | 1.21 | 1.51 | Nievergelt |
|  | Anorexia nervosa | 0.03 | 0.05 | 0.05 | 0.06 | Watson |
|  | Bulimia nervosa | 0.05 | 0.08 | 0.06 | 0.08 | – |
|  | Personality disorders | 3.00 | 4.77 | 3.91 | 4.81 | – |
|  | Cluster A | 0.09 | 0.14 | 0.11 | 0.14 | – |
|  | Cluster B | 0.95 | 1.55 | 1.19 | 1.47 | – |
|  | Cluster C | 2.04 | 3.26 | 2.75 | 3.38 | – |

### Descriptive Analysis

In the Dutch population register, we selected siblings (sharing the same legal mother and father) born between 1965 and 1985, such that they are expected to have obtained their highest diploma before the first year of diagnostic data in 2011. We obtained a final sample of 1,743,032 siblings nested within 766,514 families (**Supplementary Note** and **Supplementary Figure 1**).

Inferring the number of years of education from the final degree obtained, Dutch siblings attend education for an average of 15.35 years (SD=2.80, median=17). The sibling sample appears slightly more educated than all individuals born between 1965-1985, see **Supplementary Note** and **Supplementary Tables 1-2.**

We accessed psychiatric diagnoses based on the Diagnostic and Statistical Manual of Mental Disorders 4^th^ Education (DSM-IV) for all patients getting specialized mental care in the Netherlands between 2011 and 2016 (**Supplementary Table 3**). Specialized mental care is intended for patients with severe or complex diagnoses which require the attention of a psychiatrist or clinical psychologist. Between 2011 and 2016, the yearly incidence of psychiatric diagnoses decreased (**Supplementary Tables 4-5**). This may reflect changes in access to specialized mental health care, e.g. a 2014-reform led to an increase in the care of chronic mental health disorders by general practitioners.

Individuals can be diagnosed with more than one disorder within a year, with one primary diagnosis and one or more secondary diagnoses (**Supplementary Table 6**), or across the 6 years we studied. Psychiatric diagnoses co-occur frequently (**Supplementary Table 7, Supplementary Figure 2**) and are highly correlated (polychoric correlation up to 0.55 for MDD and PTSD). Schizophrenia is one exception as it is only weakly correlated with most other diagnoses (**Figure 1**). Genetic correlations across the GWASs selected for the MR analyses are also positive and generally substantial, but they do not always match phenotypic correlations in the Dutch population (**Supplementary Table 8**).

**Figure 1.**
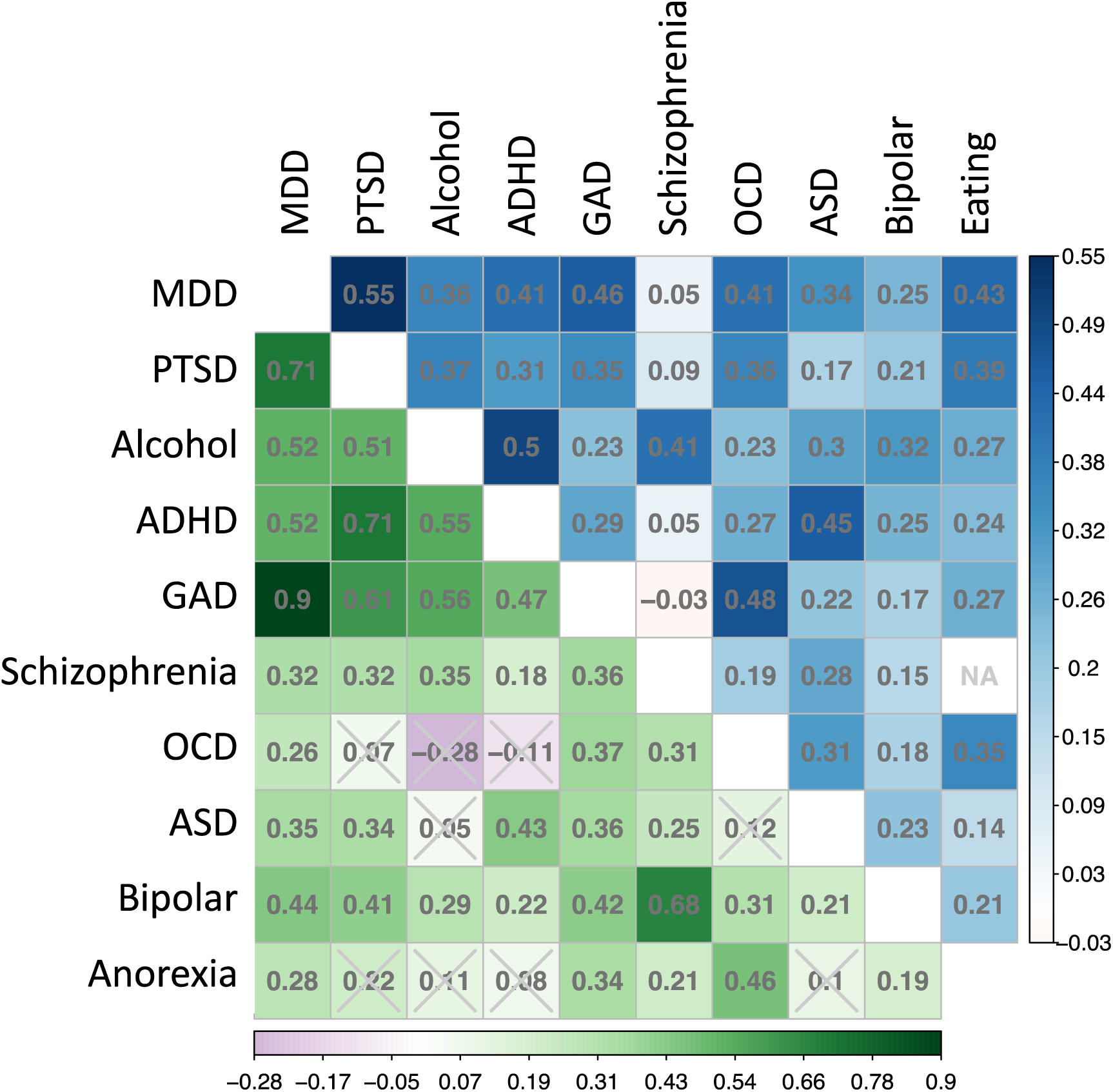
Phenotypic and genetic correlations between diagnoses. **Upper diagonal**: polychoric correlations between diagnoses in the subset of 1.7 million siblings for whom education data was available in the Dutch population registry. NA: missing value due to low cell size. In this diagonal, anorexia nervosa was combined with bulimia nervosa into an eating disorder group due to the small number of individuals for either anorexia or bulimia diagnosis. **Lower diagonal:** genetic correlations between diagnoses estimated from summary statistics of the GWASs used in the MR analyses. Genetic correlations with a two-sided pvalue higher than 0.001 (0.05/45 tests) are indicated with a gray cross. Only disorders with data available in the Dutch population register and in GWASs are represented (see **Supplementary Tables 7-8 and Supplementary Figure 2**)

**Figure 2** plots the prevalence (expressed in percentage) of each diagnosis given EA, split by sex (see also, **Supplementary Table 9, Supplementary Figure 3**). Most diagnoses have a clear sex difference in prevalence, with higher prevalence for women, except for ADHD, Alcohol-related disorders, Schizophrenia, and ASD. Most diagnoses show a decrease in prevalence with an increase in education, from 11 years to 22 years of education. The group with 11 years of education stands out in two ways. First, they started in a pre-university track (selective track) in secondary school but dropped out without obtaining a diploma or re-orienting. Second, this group has the highest prevalence for having any diagnosis, including a particularly high prevalence of bipolar disorder, schizophrenia, and ASD diagnoses. 0.75% of individuals with 11 years of education are diagnosed with bipolar disorder (below 0.4% in all other EA groups), and ∼4% of men with 11 years of education are diagnosed with schizophrenia (below 2% in further EA groups). Speculatively, dropping-out without resuming formal education could be related to prodromal symptoms for these disorders.

**Figure 2.**
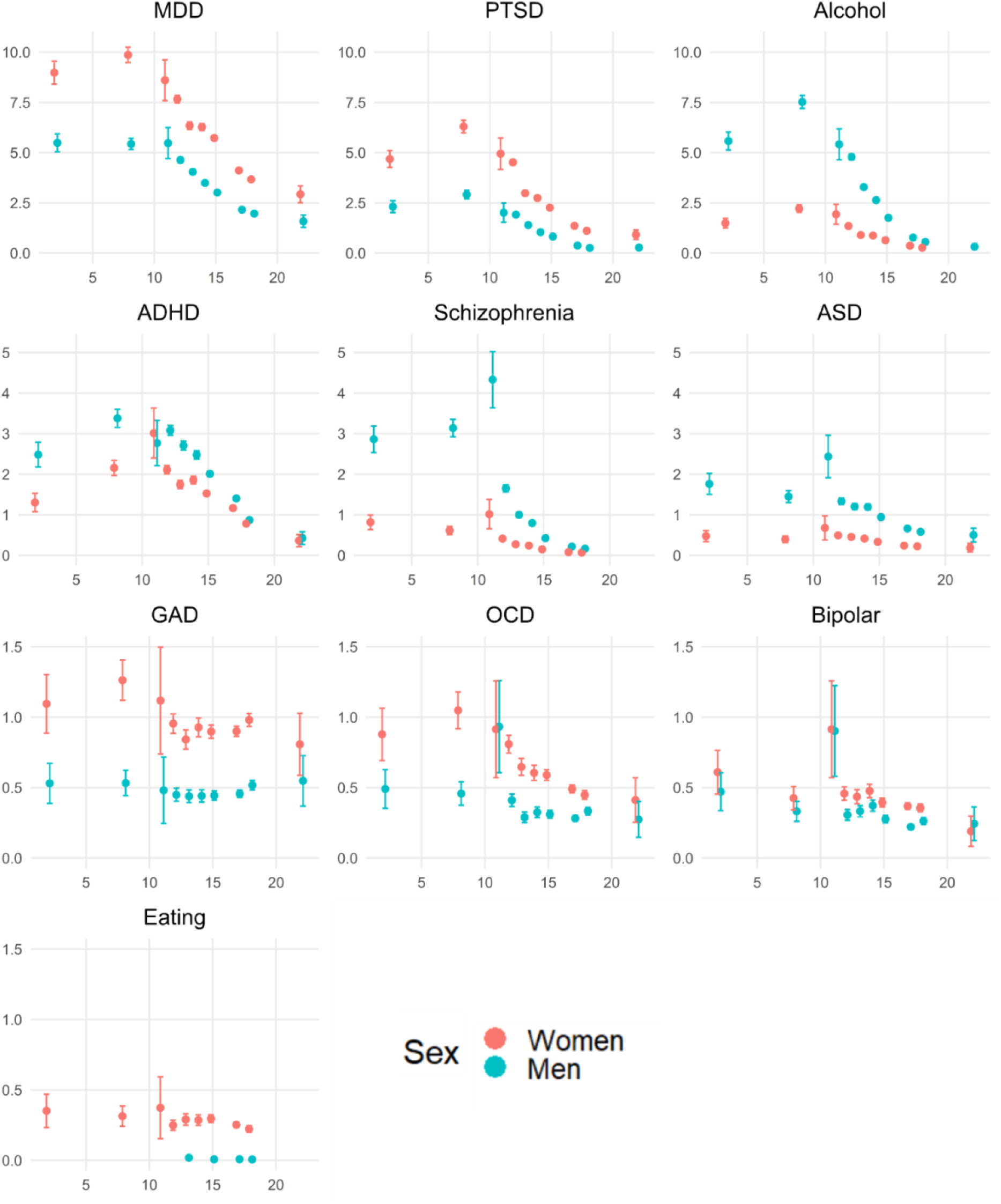
Prevalence of diagnoses given someone’s educational attainment and sex. Prevalence (expressed in percentage) of diagnoses in the sample of siblings for whom education data was available stratified by disorder (panels), number of years of education (x-axis) and sex (colour). Bars represent 95% confidence intervals. Note the scales of the y-axis are adapted depending on the diagnosis. In this figure, anorexia nervosa was combined with bulimia nervosa into an eating disorder group due to the low sample size of diagnosed individuals for some EA/sex strata. Only disorders with data available in the Dutch population register and in GWASs are represented (see **Supplementary Table 9** and **Supplementary Figure 3**)

### Within-sibship analyses

As expected, simple logistic regressions of EA on diagnoses revealed a negative association between EA and being diagnosed, for all disorders except GAD (OR=0.99, SE=0.004) (Figure 3**, Supplementary Tables 10-11**). We then ran the within-sibship regressions: we regress diagnosis status on the average EA for all siblings in a family and the deviation of the sibling’s EA from the family average. With these more robust within-sibship regressions, most associations were weaker (OR closer to 1), but still significant. On the other hand, for GAD, schizophrenia, ASD, bipolar disorder and anorexia, the within-sibship association were stronger (OR further from 1), the relationship between EA and diagnosis is stronger within than between families. Overall, odd ratios ranged from 0.98 to 0.81 per year of education, indicating a modest relation between EA and risk of diagnoses (Figure 3**, Supplementary Figure 4, Supplementary Tables 12-13)**.

**Figure 3.**
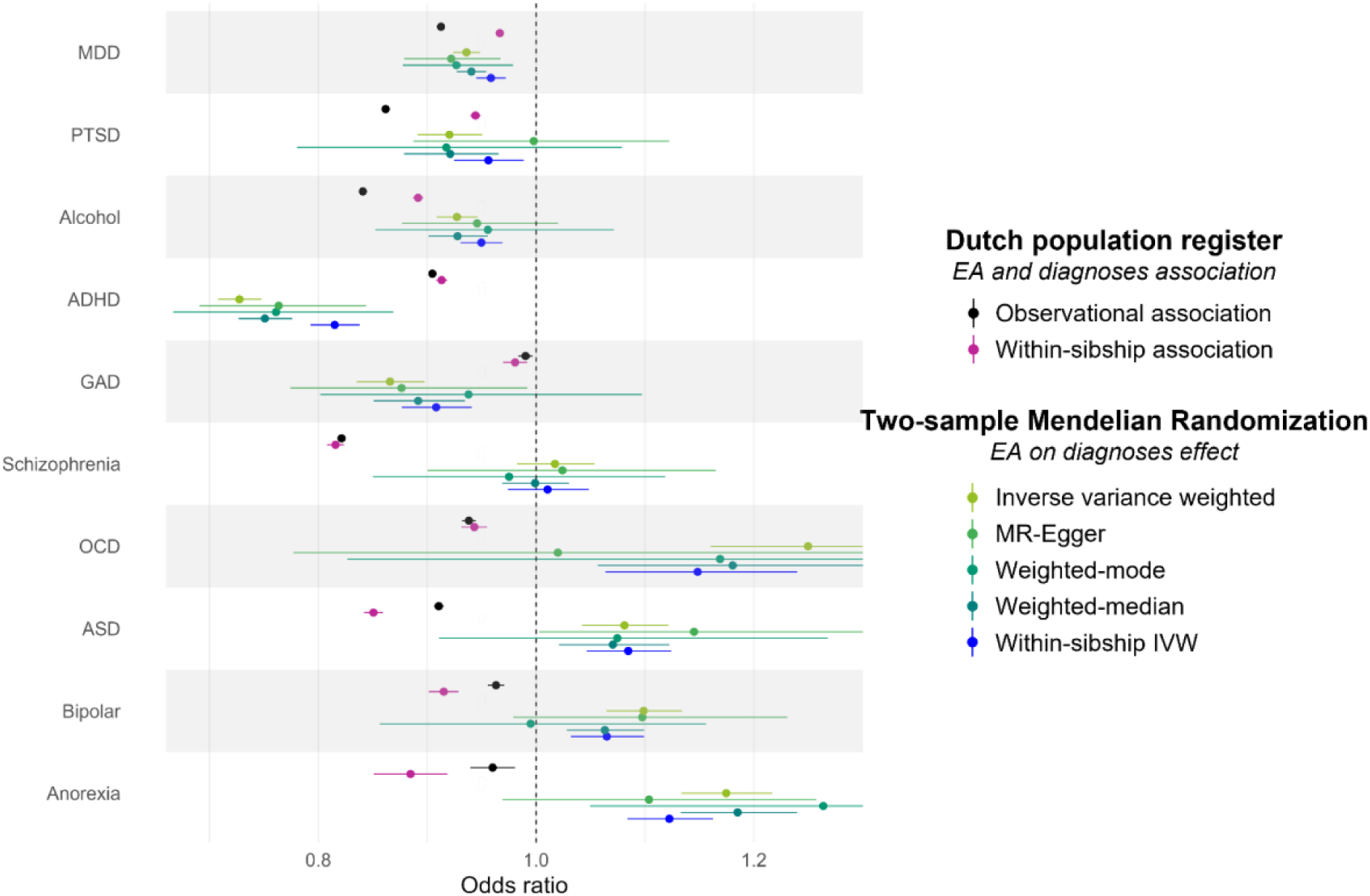
Relation between EA and diagnoses as estimated with logistic regression, within-sibship regression or MR. Odds ratios per year of education as estimated with logistic regression (black) and within-sibship models (purple) in the Dutch population register and the two-sample MR analyses of EA on diagnoses (green/blue). Bars: 95% CIs. Only disorders for which both results are available are represented (see **Supplementary Tables** and **Supplementary Figures 4 & 7**)

We run several post-hoc robustness analyses of the within-sibship regression. As for most diagnoses the prevalence differs by sex, we replicate our analysis in subsets of sibships with same-sex siblings only. For almost all diagnoses, the direction and magnitude of effects are the same. Bulimia is a notable exception: within-male sibship analysis suggests a positive relation with EA (OR=1.25, SE=0.12), but this relation is not significant (P=0.03) (**Supplementary Figure 5, Supplementary Tables 14-15**). To guard against unmodelled “loss to follow-up”, we ran analyses excluding siblings with 2 years of education (which is an implausible outcome in the Dutch system). The results are qualitatively similar, but there are subtle changes in effect sizes (**Supplementary Figure 4, Supplementary Table 16**). Estimate of the relation between EA and schizophrenia diagnosis is the most sensitive to this exclusion, with an increase of the effect of EA on schizophrenia within-sibship (OR=0.70, SE=0.005). The exclusion of siblings with 11 years of education (rare outcome of dropping-out of selective track secondary education) from our analyses did not change results (**Supplementary Figure 4, Supplementary Table 17)**.

We replicate this analysis using an alternative measure of mental care in the Dutch system: mental care expenditures, expressed in log(euro) (**Supplementary Figure 6, Supplementary Table 18**). Care expenditures include costs incurred within the specialized care, but also within the basic care (e.g. GP reporting mental health care as the reason for visit). Mental care expenditures are also negatively associated with EA (*ß*=-0.11, SE=0.00). This association is reduced within-sibship (*ß*=-0.08, SE=0.00). Comparing same-sex siblings only, the within-sibship estimate is stronger in men (-0.1) than in women (-0.06). These effects correspond to 10% and 6% decrease in expenditure per year of education within-sibship (**Supplementary Table 19**).

### Two-Sample Mendelian Randomization

#### MR estimates of EA on psychiatric diagnoses

With EA as exposure and relatively strong instruments (mean F-statistics > 50), MR IVW estimates are not always consistent with within-sibship estimates (Figure 3**, Supplementary Figure 7, Supplementary Tables 20-23**). They suggest a comparable causal effect of EA on diagnosis for MDD, PTSD, and Alcohol dependence (IVW *OR*=0.94, 0.92, and 0.93, *P* < 0.004). For ADHD and GAD, the risk-decreasing effect of EA estimated in MR is stronger than estimated in the within-sibship design (IVW *OR*=0.73 and 0.87, *P* < 0.004). Importantly, MR estimates were yet not supportive of a protective effect of EA on five diagnoses. MR shows no effect of EA on schizophrenia (IVW *OR*=1.02, *P*=0.33), and a risk-increasing effect of EA on diagnosis for OCD, ASD, bipolar disorder, and anorexia= (respectively IVW *OR*=1.25, 1.08, 1.10, and 1.17, *P* < 0.004), such that the liability to higher EA is causing a higher liability for these diagnoses. MR-Egger, weighted-mode and weighted-median estimates were mostly consistent in direction and effect sizes. Strick interpretation of evidence from pleiotropy-robust methods confirms a causal effect of EA on the liability of MDD and ADHD but not other diagnoses. I^2^ were larger than 0.9 suggesting MR-Egger’s estimates could be interpreted. There was evidence of heterogeneity for each EA-disorder pair indicating pleiotropy (Cochran’s *Q* between 821 and 2101, *Ps* < 0.004) but non-significant MR-Egger intercepts suggest the estimates are not biased by horizontal pleiotropy.

Exploratory sensitivity analyses based on SNP-effects on EA from a within-sibship GWAS yield similar or lower estimates in the same direction for all diagnoses (estimates became non-significant for PTSD) (**Supplementary Table 23)**. The reduction in effect size is particularly large for ADHD (IVW *OR*=0.73; within-sibship EA IVW OR=0.82). However, the instrument was weak (mean F-statistics ∼10.5), which can bias MR results towards the null.

#### MR estimates of psychiatric diagnoses on EA

When considering the reverse effect of mental disorder liabilities on EA, we relaxed the p-value threshold for instrument inclusion to 1e-5 for GWASs with low number of genome-wide significant hits (ASD, GAD, OCD, PTSD, alcohol dependence). The mean F-statistics for the instruments were modest (34.8 to 44), and modest-to-weak (21.5 to 22.8) when the p-value threshold was relaxed.

IVW estimates suggest a negative causal effect of the liability to disorder on education attainment (hence a bidirectional negative effect) for ADHD (IVW *ß*=-0.38, *P* < 0.004), and at p < 0.05 (a liberal threshold given the number of MR tests) for MDD (IVW *ß*=-0.32, *P*=0.010), PTSD (IVW *ß*=-0.06, *P*=0.026), and GAD (IVW *ß*=-0.09, *P*=0.016) (Figure 4**, Supplementary Tables 24-25**). We find no evidence of an effect of schizophrenia (IVW *ß*=0.00, *P*=0.94), while higher liabilities for bipolar disorder and anorexia have a positive effect on EA (IVW *ß*=0.17 and 0.26, *P* < 0.004). The estimated effects of alcohol dependence (IVW *ß=*-0.05, *P*=0.18), OCD (IVW *ß*=-0.00, *P*=0.63), and ASD liabilities (IVW *ß*=-0.03, *P*=0.40) on EA were not significant.

**Figure 4.**
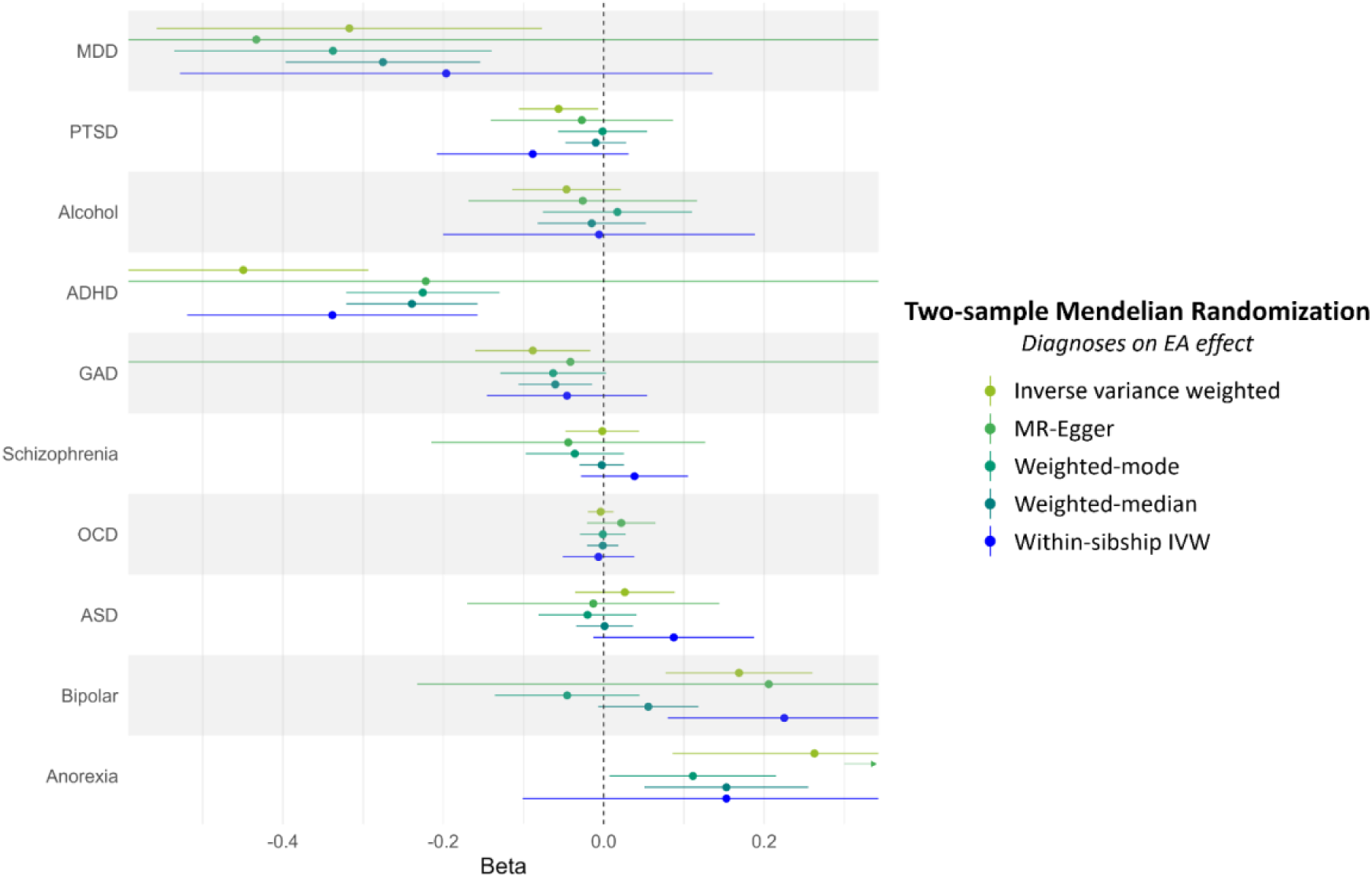
MR analyses diagnosis as exposure and educational attainment as the outcome. Effect estimates of the two-sample MR analyses of EA on diagnoses. Bars: 95% CIs. Only disorders with data available in the Dutch population register and in GWASs are represented (see **Supplementary Tables 24-25** and **Supplementary Figure 7**). The arrow in lieu of the Anorexia MR Egger highlights that the estimate is too large to be shown (*ß*=0.79, SE=0.11).

Again, MR-Egger, weighted-mode and weighted-median estimates were mostly consistent in direction and effect sizes. However, MR-Egger’s estimates had low confidence and the weighted-mode estimate was <0 (non-significant) for bipolar disorder. There was evidence of heterogeneity of effects for every disorder-EA pair (Cochran’s *Q* between 26.3 to 2602.1, *Ps* < 0.004), but for OCD-EA (Cochran’s *Q*=29.6, *P=0.04*). While the MR-Egger intercepts were never significant, the intercept for Anorexia-EA was the highest (0.06, SE=0.01), which might indicate the presence of horizontal pleiotropy. All I^2^ were larger than 0.9, however the small number of instruments for most diagnoses (e.g. Anorexia N SNPS=4) results in poor resolution to resolve directional pleiotropy. Exploratory analyses with SNP-effects based on within-sibship EA GWAS were consistent in direction and effect size with the previously described IVW estimates.

### Education of healthy siblings in the Dutch population registry

Motivated by the discordance between the within-sibship and MR analyses for bipolar disorder, anorexia, ASD, and OCD, the first implying a protective effect of education and the second a risk, we investigate the hypothesis that the (genetic) liability for being diagnosed with some disorders is in fact associated with higher education, while the disorder itself (or its prodromal manifestation) interferes with schooling. Under this model, bipolar, anorexic, ASD and OCD patients are expected to have a higher familial and genetic liability for EA than the general population. Therefore we expect the healthy siblings of these patients, but not of other patients, to be more highly educated than individuals in unaffected families. In the Dutch register, we do find that siblings of patients have less education than siblings in unaffected families for all disorders but bipolar disorder and anorexia (Figure 5**, Supplementary Table 26**). Unaffected siblings of patients with bipolar, ASD and anorexia have the same average years of education as unaffected families (average EA of unaffected families = 15.54; bipolar = 15.58, ASD = 15.58, anorexia = 15.75, t-test P=0.21 for bipolar, 0.11 for ASD, 0.005 for anorexia). Siblings of Bipolar II patients had slightly higher EA than individuals in unaffected families (15.69, p <0.004) (**Supplementary Figure 8**).

**Figure 5.**
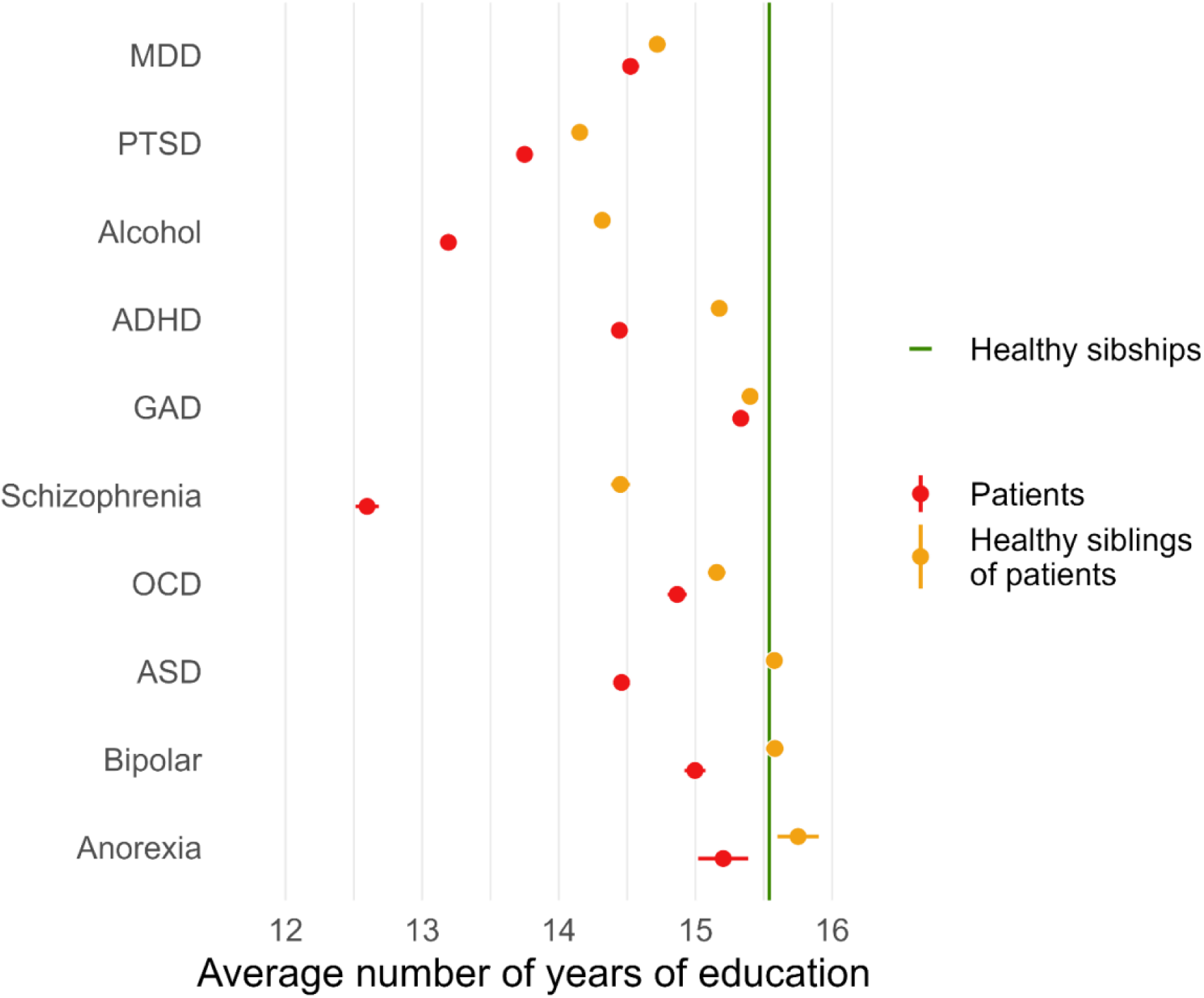
Average number of years of education of patients, of healthy siblings of patients and unaffected sibships. The green line is the average number of years of education of unaffected sibships (15.54 years of education), in families in which none of the siblings was diagnosed with a mental disorder between 2011 and 2016. Red dots are the mean education of affected siblings per diagnosis. Orange dots are the mean education of siblings of an affected sibling that are themselves were never diagnosed in the 2011-2016 timeframe. Bars: 95% CIs. Only disorders with data available in the Dutch population register and in GWASs are represented (see **Supplementary Table 26** and **Supplementary Figure 8**).

## Discussion

Triangulating across two designs, we find consistent protective effects where higher EA reduces the risk of MDD, alcohol dependence, GAD, ADHD, and PTSD diagnoses, though for some traits, there is also evidence for reverse causation. Critically, for MDD, alcohol dependence and PTSD, the implied causal effects in MR and within-sibship designs are substantially lower than the observational association. For other diagnoses, notably bipolar disorder, anorexia, ASD and OCD, the two designs yield inconsistent results. These are noteworthy and gave rise to additional analyses. Our study reveals that statistically convincing findings based on either design would have provided an incomplete or misleading understanding of the relationship between education and mental health. We discuss the findings for some common diagnoses, highlighting representative results. While we center our discussion on MDD, ADHD, schizophrenia and bipolar disorder, we speculate similar interpretations could be extended to disorders with similar patterns of results.

### Depression

Both the within-sibship and the MR approach suggest around 6% lower odds of depression diagnosis per additional year of education, while there is also suggestive evidence for reverse causation (depression diagnosis or liability hampers EA). The implied causal effect of education on the risk of depression diagnosis is lower that the observational relationship. Our results are consistent with work that relies on other quasi-experimental methods, for example exploiting compulsory school-law reforms lengthening minimum EA^32^. One study found a ±7% reduction in the probability of meeting diagnostic criteria for depression (*N*=3,704)^33^. Another study (*N*=21,085) found no significant effect of EA on depression, though their negative sign is consistent with our findings^34,35^. Multiple previous (MR) studies based on UK Biobank data, which is an older (age 40-70) and more highly educated cohort, find no effect^34,36^. The inconsistency with the previous literature might be due to their samples of healthy volunteers, while our within-sibship sample is population-based. An additional year of education might especially benefit the mental health of those at the lower end of the education distribution (a group under-represented in volunteer samples)^32^. Also, the protective effect of EA on mental health might wane with age^19^.

We focussed on individual’s final obtained degree. However, EA isn’t the only aspect of education that relates to mental health. For example, a German reform reduced the academic high-school track by one year while keeping the curriculum constant. This intensified schooling increased depressive symptoms^37^. Others have also shown that kids being relatively young for their school grade are more likely to be diagnosed with depression^38^ or that later separation of students into different tracks might increase depression in women^39^. While we find that higher EA plausibly reduces the risk of many diagnoses in adulthood, any reform aimed at increasing EA should keep in mind that other aspects of education might have opposite effects.

### ADHD

For ADHD reciprocal processes are plausible. The formal diagnosis of ADHD alludes to problems that interfere with school work. This creates an obvious dependence between ADHD and poor performance in school (socially or academically). Other two-sample MR studies also reported the bidirectional effect of EA and ADHD. Additionally, they suggest this effect is independent of IQ^27,40^. Interestingly, our MR estimate of the effect of EA on ADHD is lower when using SNP-effects based on within-sibship EA GWAS, suggesting that assortative mating, population stratification, and/or gene-environment correlation might bias MR estimates. These biases could explain the difference in estimates between MR and the within-sibship analysis. Besides, the difference between the observational and within-sibship association registry is small, suggesting familial factors do not explain the association between EA and ADHD diagnosis.

ADHD is part of the “disorders usually first diagnosed in infancy, childhood, or adolescence” in the DSM-IV, hence usually diagnosed at school age, which seem inconsistent with a causal effect of EA on ADHD diagnoses. Our data concerns ADHD diagnoses in adulthood. As we do not have data on age at first diagnosis, we cannot distinguish between a recurring diagnosis and first diagnosis. However, there are plausible mechanisms through which EA might continue to affect the likelihood of diagnosis after one’s educational trajectory. For example, higher-educated individuals might better cope with their ADHD symptoms and might have more power to select suitable environments (e.g., selection of jobs with more flexibility^41^).

### Schizophrenia

The within-sibship design estimates 18% lower odd of schizophrenia per additional year of education, while MR suggests no causal relationship. The within-sibship association might reflect the interfering effect of prodromal or early symptoms of schizophrenia on education. This would be consistent with our observation that 4% of Dutch men who drop out of the pre-university high school track, and do not re-enter education, are diagnosed with schizophrenia. However, we do not observe a causal effect of schizophrenia on EA in our MR analysis. Note that the interpretation of MR is difficult when the exposure is binary and rare. The MR estimates should be viewed as reflecting the effect of the liability for being diagnosed, not an effect of the diagnosis itself. It is easy to imagine how core symptoms of schizophrenia, like delusions, hallucinations, and disorganized thoughts, interfere with education, but these symptoms are not gradually experienced over the entire liability spectrum. If we do accept the MR findings, the discrepancy between the within-sibship and the MR results could be alternatively explained by factors not shared between siblings that reduce EA and increase the risk of schizophrenia diagnosis (e.g. risk exposures such as trauma). Contrasting our findings, previous MR estimates using earlier schizophrenia GWASs hint at an increased risk for schizophrenia per additional year of education^26,28^, as we found for anorexia, OCD, and bipolar disorder.

### Bipolar disorder, anorexia nervosa, ASD, and OCD

Bipolar disorder, anorexia nervosa, ASD, and OCD have the most striking pattern of results. While the within-sibship design suggests that each additional year of education *reduces* the likelihood of being diagnosed by 6 to 12%, the MR estimates suggest that EA *increases* the likelihood. We hypothesised that these diagnoses follow a model where the (genetic) liability for being diagnosed is associated with higher education, while the disorder itself (or its prodromal manifestation) interferes with schooling. Supporting this, we found (1) a high prevalence of bipolar disorder among pupils dropping out of the pre-university high-school track and (2) healthy siblings of bipolar, anorexic and ASD patients have similar or even higher EA than the average unaffected sibships.

Other Dutch studies comparing siblings of patients confirm this observation for bipolar disorder compared to schizophrenia or MDD^42,43^. However, this was not replicated in Denmark^44^. Bipolar disorder is associated with traits that are valuable in school settings like more creativity^45,46^ and higher childhood IQ^47^. A Swedish population-cohort study reports that individuals with excellent high-school performance have a fourfold increased risk of bipolar disorder over those with average grades^48^. Similar associations with higher education of family members, higher school grades and higher IQ is found for anorexia^49–51^, patterns are more unclear for ASD^52,53^ and OCD^54,55^.

A true positive relation between factors that increase both success in education and risk of bipolar and anorexia diagnoses seems supported, although the mechanism remains unclear. They could share biological mechanisms or psychological traits like creativity. Likewise, higher-educated individuals, or individuals coming from higher-educated families, could be more likely to be diagnosed with these disorders, due to more proactive help seeking, better access to care that facilitates these diagnoses, or preferential diagnosis by practitioners. Finally, biased selection into GWASs could result in the selection of patients with higher-than-average education.Further studies are essential to understanding the mechanisms of the apparently contradictory association between these disorders and EA.

### Limitations

Our conclusions rely on within-sibship and MR estimates, and on whether we can interpret contradictory findings. We assume these contradictions are mainly due to true differences in underlying phenomena. However, other statistical, measurement-, and sampling-related factors could play a role.

The two methods rely on data obtained in different samples drawn from different populations. Our within-sibship analysis was based on registry data, containing most of the Dutch population born between 1985-1965, so highly representative for the Netherlands. In contrast, the MR is based on GWASs of international samples who are volunteers of European-ancestry. Both higher risk for mental illness and lower EA are known to increase non-participation and participant attrition^56–59^, a selection bias that could induce a collider bias^60^. In the within-sibship analysis, we rely on diagnoses by professionals in specialized care, therefore dependent on the current referral policies of the Dutch healthcare system. All our GWASs rely on diagnoses, but they differ on the report (self-report, medical files, etc.) and on the timeframe (most look at lifetime diagnoses, but the age range of participants vary widely). In both types of data, who seeks help and who is diagnosed may depend on class, ethnicity and context. Our conclusions are phrased in terms of diagnoses; potential mechanisms discussed are speculative. Any conclusions beyond diagnoses run into the imperfect relation between people’s symptoms and people’s DSM-IV diagnosis. Within a national register, undetected “loss to follow-up” resulting from death or migration might bias results. However, when conducting sensitivity analyses that excluded individuals with educational durations inconsistent with Dutch education laws (which could indicate death or migration before completing education), the results remained largely unchanged.

Regarding two-sample MR, we use MR analyses robust to weak-instrument and pleiotropy, but the additional statistical power required make the estimates more uncertain. To control for potential biases due to demographic and dynastic effects, we used summary statistics from a within-sibship GWAS. However, EA is the only trait with a within-sibship GWAS suitable for MR, leading to an unbalanced control of these biases in our MR. Notably, dynastic effects could lead to false positive bidirectional effects^61^. Similar well-understood caveats apply to sibling designs but most additional criticisms of this design^8,9^ pertain to lack of insight on the direction of effect and the possible underestimation of effect sizes.

Further insights might be gained by using additional independent causal inference techniques, such as comparing diagnostic rates among individuals who left formal schooling immediately before or after the implementation of educational policies that changed the duration of schooling^35,36^.

Finally, our findings pertain to the effects of education on mental health within the current confines of social life and social policy. Our findings do not identify an immutable or permanent cause of differences in mental health. Welfare policy changes not directly related to education (e.g. minimum wage, affordable living, quality housing) may improve people’s mental health directly and could as effectively close the health gap between educational groups as educational policy changes could.

### Conclusion

The aforementioned caveats limit the certainty we should ascribe to specific causal claims. Nevertheless, we established consistent potential causal effects of EA on the risk of being diagnosed with MDD, ADHD, PTSD, alcohol dependence, and GAD. Except for GAD, these potentially causal effects are smaller than the observational associations. For diagnoses like bipolar disorder and anorexia, our results suggest a positive relationship between EA and the diagnosis liability, yet a negative relationship between EA and the diagnosis itself. These patterns deserve further study and would have been missed when applying a single causal-inference technique in isolation.

## Methods

This study was pre-registered: https://osf.io/vmpfg/?view_only=b17c64b5600c4d32902e55ea26d63f37. Deviations to the preregistration are detailed in **Supplementary Note**. All code associated with the analyses is available on GitHub at https://github.com/PerlineDemange/CBS-MR. We follow the STROBE^62^ and STROBE-MR^63^ reporting guidelines (**Supplementary Tables 27-28**). This research was reviewed and approved by the Scientific and Ethical Review Board (VCWE) of the Faculty of Behaviour & Movement Sciences, VU University Amsterdam; application number VCWE-2020-054.

## Within-sibship design

## Data source

We analyze restricted access microdata from Statistics Netherlands (CBS). Under strict conditions, these microdata are accessible for statistical and scientific research. For further information on remote access procedures.

## Study population

We included individuals born between 1965 and 1985 (N=6,539,767), such that they are between 26 and 46 years old when the first year of diagnostic data is available. From these we select siblings (sharing the same legal mother and father), where more than one sibling has educational data available. We retain a final sample of N=1,743,032 individuals nested within 766,514 families. For a comprehensive overview of the selection procedure, see **Supplementary Note** and **Supplementary Figure 1.**

## Educational attainment

Educational attainment data is based on various registers and surveys and has a high coverage (more than 11 million people). Based on the final degree obtained we inferred the number of years of full-time education of the individual. The transformation of the 17 diploma categories to years of education is available in **Supplementary Table 1**. For a comprehensive overview of the variable definition, see **Supplementary Note.**

## Mental health outcomes

The **Dutch mental health care system** distinguishes two systems of care. Here we rely on diagnostic data for specialized/second-line care. Specialized mental care is intended for patients with severe or complex diagnoses which require the attention of a psychiatrist or clinical psychologist. **Psychiatric diagnoses** are obtained from the care trajectory of patients getting specialized mental care. The **Supplementary Note** provides further explanation of the Dutch mental health care system. Diagnoses are classified based on the Diagnostic and Statistical Manual of Mental Disorders 4^th^ edition (DSM IV). We consider an individual as affected if they were diagnosed with any of the disorders listed in **Table 1**, in at least one year during the 2011 to 2016 period. The diagnosis and subdiagnosis categorization is reported in **Supplementary Table 3**. Regarding ASD, we consider a definition closer to the DSM V (spectrum), also used for the GWAS, including patients with: autistic disorder, childhood disintegrative disorder, Asperger syndrome, and pervasive developmental disorders not-otherwise-specified.

**Mental health care expenditures**^64^ are assessed in two ways: expenditures from the first line/basic care and expenditures from the 2nd line/specialized care. We summed basic and specialized care for each year. Due to the steeply skewed distribution of incurred mental health expenditures (**Supplementary Table 18, Supplementary Figure 6**), we averaged the expenditures across 2009-2018 for each individual and log-transformed the personal average.

## Statistical analyses

All diagnoses analyses were done in the sibling sample (4), for each mental health diagnosis separately. We estimate polychoric correlations between all diagnoses. We ran an observational analysis: a logistic regression with EA as a predictor and the psychiatric diagnosis as outcome, ignoring family structure. We then ran within-sibship logistic regression analysis. We regress diagnosis status on the average EA for all siblings in a family and the individual deviation of the sibling’s EA from their family average. The effect of the average EA (between-sibship effect) represents the expected change in the outcome being diagnosed given a one-unit (the unit being scaled as a year of education) change in the sibling average, while the effect of the deviation of the sibling to their family average (within-sibship effect) represents the effect of EA when keeping the factors common to the family constant. We correct standard errors for family clustering using lmtest and sandwich R packages in these analyses. For the analysis of mental health care expenditures, we fit a linear model instead of a logistic model as the outcome is continuous, and also report results from linear models with random family effects. In all analyses, we included sex, birth year and birth order as covariates.

As sensitivity analyses, we separately considered men and women (from same-sex sibships) and we omitted specific education groups that are rare or implausible given the Dutch educational system (11 and 2 years of education). We investigated mean differences in EA between patients, siblings of patients, and families that are entirely unaffected. We performed t-test to compare the mean EA of these groups for each disorder. All statistical tests are two-sided.

## Two-sample Mendelian Randomization

We follow recommendations by Burgess et al^65^.

## Summary Statistics

We relied on summarized statistics from a large well powered 2018 GWAS of EA^13^ (EA3). We reproduced this GWAS by meta-analyzing published summary statistics with summary statistics obtained from 23anMe, Inc, as done in ^66^. For additional sensitivity analyses, we relied on summary data from the within-sibship GWAS of EA^17^. The within-sibship GWAS is significantly smaller, but because the SNP-effects are estimated within-family they are unbiased by potential effects of assortative mating, population stratification or intergenerational genetic effects^67,68^.

For each psychiatric disorder, we selected GWAS summary statistics preferentially selecting the most recent or largest GWAS available. A full list of GWASs and a description of the summary statistics are available in **Supplementary Table 20**. We assessed potential sample overlap between EA and psychiatric disorders GWASs using LD-score cross-trait intercept as proxy^69^ (**Supplementary Table 21** & **Supplementary Note**).

## Choice of the genetic variants

For data cleaning and analyses we used TwoSampleMR^70^ in R.4.1.0^71^. When analysing EA as the exposure, we first excluded genetic variants not present in the outcome summary data. For EA3 summary statistics, we selected genetic variants associated with EA at p < 5e-8. For within-sibship EA summary statistics, we selected significant independent loci identified by the EA3 GWAS, which were also associated at p-value <0.05 in the within-sibship EA GWAS. We then clumped to select statistically independent variants (kb=1000, r^2^=0.001). When analysing psychiatric disorders as the exposure, if clumping genetic variants associated at p < 5e-8 led to the selection of less than 5 genetics variants, we further selected genetic variants associated with the exposure at p<1e-5 (this occurred for ASD, GAD, bipolar-II disorder, OCD, PTSD, and alcohol dependence). Variants were then harmonized between exposure and outcome summary statistics, ensuring the SNP effect relates to the same reference allele. Ambiguous and palindromic variants with MAF > 0.42 were excluded.

We scaled the effect sizes from the two EA GWASs so that the SNP-effects reflect change in term of years of education. For this, we estimated the weighted average standard deviation of the education phenotype in the cohorts included in the EA3 study (SD=3.9 years) and multiplicated both SNP-effects and their SEs by this number^27,72^. Estimates for the psychiatric disorders SNPs were converted to log(OR) if reported in OR. MR estimates were transformed back to the OR scale where needed.

## Analyses

We ran two sets of MR analyses: EA as exposure and psychiatric diagnoses as an outcome, and psychiatric diagnoses as exposure and EA as an outcome. We ran an inverse-variant weighted (IVW) mendelian randomization^12^. We judged the significance of the p-value following a Bonferroni correction: significance threshold 0.05/12 traits = 0.004. To test for the robustness of the IVW findings against potential violation of the MR assumptions, we ran MR-Egger^14^, weighted-mode^16^, and weighted-median^15^ analysis. We reported the Cochran Q’s-statistic SNP effect heterogeneity and the F-statistics assessing potential weak instruments bias^73,74^. Additionally, we reported the I2 statistic^75^, which gives an indication of the violation of the NO Measurement Error (NOME) assumption, on which MR-Egger relies. We also report LD-score based genetic correlations between all GWASs, computed with genomic SEM. All statistical tests are two-sided.

## Code and Data availability

All code associated with the analyses is available on GitHub at https://github.com/PerlineDemange/CBS-MR. We analyze restricted access microdata from Statistics Netherlands (CBS). Under strict conditions, these microdata are accessible for statistical and scientific research. For further information on remote access procedures. For GWAS summary statistics availability, see original publications. For 23andMe, Inc. dataset access, see https://research.23andme.com/dataset-access.

## Contributions

P.A.D. and M.G.N. conceived and designed the study, with helpful suggestions from E.v.B. and D.I.B. P.A.D. analysed the data, with support from M.G.N. for the MR analyses. P.A.D. designed the figures and drafted the manuscript. All authors contributed to and approved the final version of the manuscript.

## Data Availability

We analyze restricted access microdata from Statistics Netherlands (CBS). Under strict conditions, these microdata are accessible for statistical and scientific research. For further information on remote access procedures. For GWAS summary statistics availability, see original publications. For 23andMe, Inc. dataset access, see https://research.23andme.com/dataset-access.

## Acknowledgments

We thank the Open Data Infrastructure for Social Science and Economic Innovation (ODISSEI: https://ror.org/03m8v6t10) for financing access to Statistics Netherlands microdata via a microdata access grant awarded to P.A.D. and a member discount. P.A.D. is supported by the grant 531003014 from The Netherlands Organisation for Health Research and Development (ZonMW). P.A.D. is funded by the European Union (Grant agreement No. 101045526). Views and opinions expressed are, however, those of the authors only and do not necessarily reflect those of the European Union or the European Research Council Executive Agency. Neither the European Union nor the granting authority can be held responsible for them.D.I.B. is supported by the Royal Netherlands Academy of Science (KNAW) Professor Award (PAH/6635). E.v.B. is supported by ZonMW grant 531003014 and VENI grant 451-15-017. M.G.N. is supported by R01MH120219, ZonMW grants 849200011 and 531003014 from The Netherlands Organisation for Health Research and Development, a VENI grant awarded by NWO (VI.Veni.191 G.030) and is a Jacobs Foundation Research Fellow. We thank the research participants and employees of 23andMe, Inc. for making this work possible.

