## Supplementary Notes and Figures for "Evaluating the causal relationship between educational attainment and mental health"

### Supplementary Information

### Supplementary Note

#### Supplementary Methods

##### Deviations from preregistration

We deviate from the preregistration in few elements of the analyses. Justifications for these changes is reported in Methods and Supplementary Methods paragraphs below.

- For the within-sibling analyses:
    - We excluded individuals who have special needs education as highest degree obtained.
    - We did not run analyses for the diagnoses oppositional defiance disorder and conduct disorder because of excessively low case numbers.
    - We averaged the expenditures across 2009-2018 for each individual and log-transformed the personal average, due to skewed distribution, but also to enable us to express the effect in % change in expenditure per year of education.
    - We ran exploratory sensitivity analyses: within same-sex sibships only, excluding individuals with 2 years of education, and excluding individuals with 11 years of education
    - We adjusted standard errors for family clustering.
- For the MR analyses:
    - We did not look for proxy SNPs when the sentinel SNP was absent, rather we first selected the SNPs present in both GWAS and pruned lead SNPs from that intersection (ensuring substantial overlap).
    - We scaled the SNP effects on EA based on the standard deviation of the number of years of education in the cohorts present in the GWAS (SD=3.9).
    - We ran exploratory sensitivity analyses: MR analyses with within-sibship EA SNP effects.

##### Within-sibling design

*Selection of the study population within the Dutch national registry*

We selected individuals born between 1965 and 1985 (N = 6,539,767), such that they are between 26 and 46 years old when the first year of diagnostic data is available and they are reasonably expected to have completed their full-time education (sample (1)). We selected individuals with available parental information (N = 4,063,765) to identify sibling pairs. Parental information was based on a municipal personal record database combined with linked parent-child data. We defined full siblings as individuals sharing the same legal mother and father (total N = 3,282,626). We excluded families whose parents had more than one child with another partner (two or more sibships with one parent in common) to avoid dependence between the selected sibships (N = 3,254,901). We cannot ensure our population constitutes biological siblings only, but we performed the following additional exclusions to remove siblings unlikely to be biological siblings. We excluded siblings with same-sex parents (N = 293). Some families had mismatching parental information: while the parental identifier was identical for both children, the reported parental birthday or country of birth was different. We excluded individuals for whom this parental information was inconsistent and removed sibships with only one child left (N = 17,293). While keeping siblings born on the same date (probable twins/multiples, N = 76,672), we excluded siblings born less than 6 months apart and removed sibships with only one child left after applying the filter (N = 2,392). We computed the birth order for all siblings (giving twins and multiples the same rank). These siblings constitute sample (3).

Individuals with missing education were excluded, as well as families where only one child had education (we retain 1,743,826 individuals in 766,832 families). We excluded individuals whose higher diploma was special needs education (referred to as *Praktijkonderwijs* in Dutch*)* and any siblings who then had no further siblings in the data (N = 794). We retain a final sample of N = 1,743,032 individuals nested within 766,514 families (sample (4)). The flowchart of the study sample selection is presented in **Supplementary Figure 1**. For descriptive comparison purposes, we excluded individuals from sample (1) that had no educational data or whose highest diploma was special needs education to create sample (2) with N = 3,305,733.

Mental health diagnoses were available for all as we assume no available record equals no diagnoses (N=1,743,032), while mental health care costs were available for 1,688,353 individuals part of a complete sibship with EA data (sample (5)).

For sensitivity analyses, we considered complete sibships with only same-sex siblings, as well as complete sibships excluding individuals whose number of years of education is 2 or 11.

##### *Educational attainment*

Educational attainment data is based on various registers and surveys. Unlike other CBS datasets, this dataset doesn't cover the entire Dutch population but has very high coverage (more than 11 million people). Based on the final degree obtained we inferred the number of years of full-time education of the individual. For degrees that ambiguously map to different numbers of years of education, we pick the lowest (e.g. master's at university can be one or two years in the NL, we selected 1 year). Individuals whose highest obtained education is special needs education (*Praktijkonderwijs)* are rare (less than 0.05% of the population). While most other special needs education students go on to complete additional practical diplomas, these students attended special needs education since they left primary education until they are at least 18 years old. As a result, we omit these individuals from the analysis. The transformation of the 17 diploma categories to years of education is available in **Supplementary Table 1**.

##### *Mental health outcomes*

The **Dutch mental health care system** (Geestelijke Gezondheidszorg GGZ) is split into two systems of care, which were until 2014 referred to as first-line care and second-line care, and since referred to as basic and specialized care. The reform in 2014 changed what types of mental health problems are treated in general medicine, basic psychological care and specialized care. As a rule of thumb, general practitioners treat simple problems, which do not require a DSM disorder diagnosis or specialized interventions. In basic mental care, patients with a single well-defined DSM disorder that are manageable in an outpatient setting are treated. Basic psychological care is limited in duration and cannot exceed 8 sessions. Beyond these sessions, care is provided by general medicine if mental health is stabilized, or by specialized care, if further mental health care is needed. Specialized mental care is intended for patients with severe or complex diagnoses which require the attention of a psychiatrist or clinical psychologist (in both inpatient or outpatient settings). When entering specialized care, a “Diagnose Behandeling Combinatie (DBC)” is registered, describing the primary diagnosis of the patient, potential additional secondary diagnoses, and the treatment plan (for a maximum of a year). This last group of diagnosis are included in our current diagnoses analysis. Patients with long-withstanding disorders receive a DBC only if they are treated in specialized care. If their mental health is stabilized, they will not have a DBC. Diagnoses reported in the DBC are only diagnoses pertaining to the current mental health problems of the patient: for example if a patient was diagnosed as a child with ADHD, but this diagnosis does not fit current symptoms, ADHD will not be reported.

**Psychiatric diagnoses** are obtained from the DBC care trajectory of patients getting specialized mental care. Diagnoses are classified based on the Diagnostic and Statistical Manual of Mental Disorders 4$^{th}$ edition (DSM IV).

Diagnoses are available per year and are binary: the individual either has been diagnosed or not. An individual is considered diagnosed in our analysis if they were diagnosed with any of the disorders listed in **Table 1**, in at least one year during the 2011 to 2016 period. Due to the low sample size (N=174 and 98 respectively in sample (4)), conduct disorder and oppositional-defiant disorder diagnoses were not studied, unlike preregistered.

**Mental health care expenditures** are split over two variables: expenditures from the first line/basic care and expenditures from the second line/specialized care. These expenditures include expenditures reimbursed by mandatory health insurance. Due to long waiting lists for mental care in many areas, patients might look for temporary psychological help without a GP referral, and with professionals whose expenditures might only be partially covered by mandatory insurance. It does not include expenditures covered by supplementary private insurance. The expenditures reported in our data can therefore be an underestimation of the total expenditures incurred by a given individual.

We summed basic and specialized care for each year. As care expenditures changed in 2014 due to healthcare reform, we treated pre- and post- 2014 separately for missing data and exclude individuals who had less than 50% of the years available between 2009 and 2013 and/or between 2014-2018. We exclude individuals with negative expenditures reported. We deviated from the planned transformation of variables (standardization and average of the two-time ranges), due to the steeply skewed distribution of incurred mental health expenditures (**Supplementary Table 18, Supplementary Figure 6**). Rather, we averaged the expenditures across 2009-2018 for each individual and log-transformed the personal average. We then excluded individuals without education data and without siblings with complete data.

##### *Sensitivity analyses*

As sensitivity analyses designed to detect possible sex difference, we repeated the regressions described in Methods in same-sex sibships. We additionally repeated the analyses in a subsample excluding siblings with 11 years of education (which corresponds to dropout from university-track secondary education and is associated with an abnormally high risk of some diagnoses) and individuals with 2 years of education (an implausible educational duration under Dutch education laws).

#### Two-sample Mendelian Randomization

##### *Summary statistics for psychiatric disorders*

For each psychiatric disorder, we included summary statistics obtained from the most recently performed GWAS or with the biggest sample size. Well-powered GWASs, with $h_2$ z-score > 2, are only available for a few psychiatric disorders. A full list of GWASs and a basic description of the summary statistics are available in **Supplementary Table 20**. All GWASs were performed in European-ancestry participants only.

GWASs of EA and a few GWASs of the psychiatric disorder include some overlapping samples. Overlap can strongly bias the MR estimates in a two-sample analysis, direction and size of the bias varying depending on the degree of overlap (Burgess et al., 2016). To identify potential overlap, we measured the LD score cross-trait intercept (Bulik-Sullivan et al., 2015) using the ldsc function of the

GenomicSEM R package (Grotzinger et al., 2019). When the cross-trait intercept was significant ($p < 0.05$), we further investigated potential overlapping cohorts (**Supplementary Table 21**).

##### *Investigation of the sample overlap in GWAS*

Most GWASs had a low sample overlap with the EA GWAS (cross-trait intercept non-significant **Supplementary Table 21**). PTSD GWAS seems to share sample with EA GWAS: the PTSD GWAS contains UK Biobank participants. However, the (Nievergelt et al., 2019) PTSD GWAS is the only well-powered PTSD GWAS with summary statistics available, so we still used these summary statistics in our MR analyses.

#### Comparison of the population and the sibling subsample

Statistics Netherlands allows to access information for the entire registered population of the Netherlands. Sibships born between 1965 and 1985 are a non-random subset of this population. Our siblings subset reports significantly higher education than the total population (mean years of education 15. 35 versus 14.74, median 17 versus 15) (**Supplementary Table 2**).

Regarding mental health diagnosed, the population prevalence is an underestimate of the true population prevalence due to missing data being treated as non-diagnosed individual. We expect sample with EA data available to be closer to the true estimate, as this likely is more representative of individuals who stably live in the Netherlands. As such we see that our sample with available EA data has an higher rate of psychiatric diagnoses. Comparing the total population with EA and our siblings subset, the prevalence for psychiatric diagnoses between 2011-2016 is similar (0.1094 for the total population, 0.1057 for the siblings subset). Prevalence and sample sizes over years and over psychiatric diagnoses for each sample group are available in **Supplementary Tables 4 & 5**.

### Supplementary Figures

#### Supplementary Figure 1. Flowchart of study sample

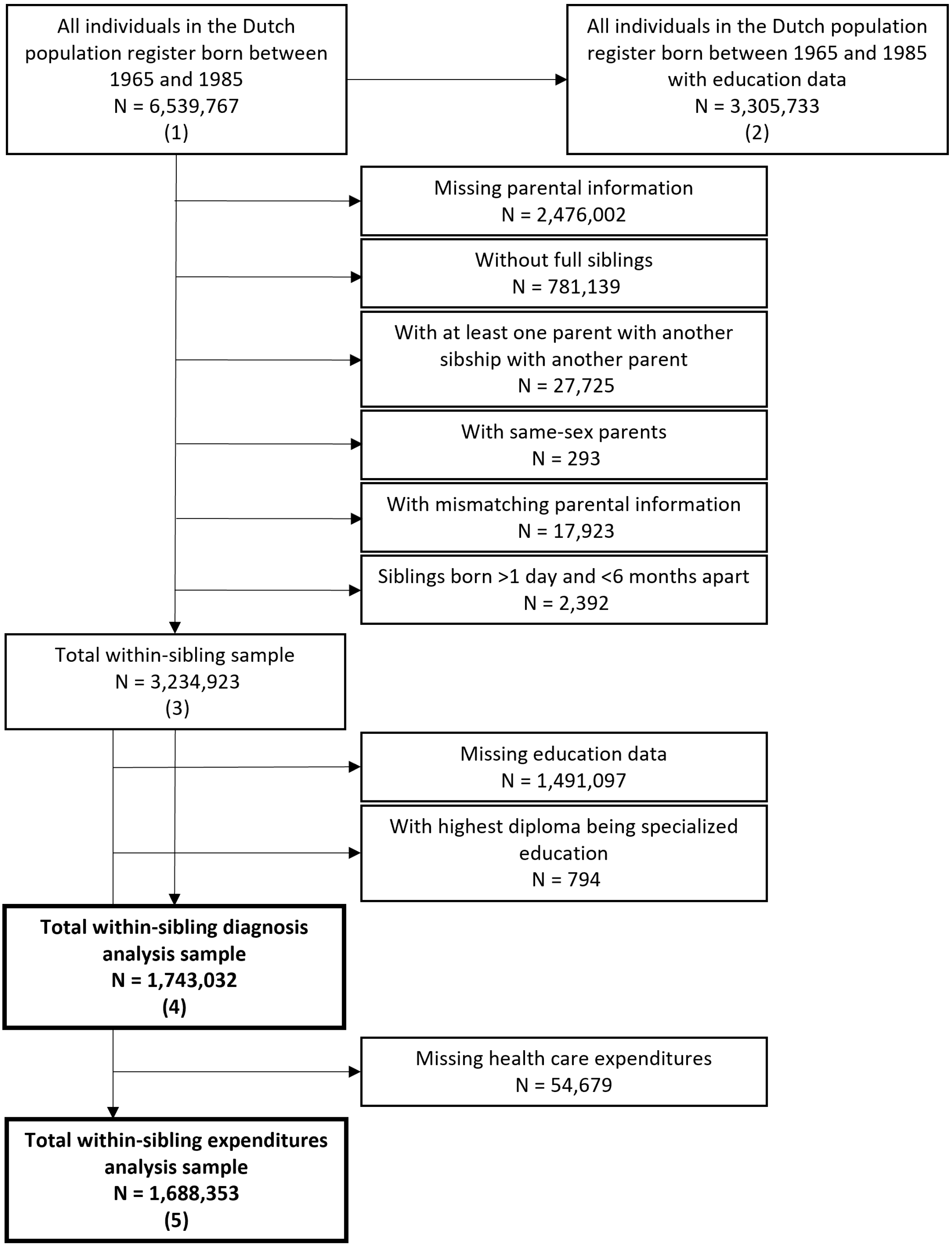

#### Supplementary Figure 2. Cooccurrence, polychoric correlations and genetic correlations of diagnoses

**A. Cooccurrence of diagnoses**

Color-coded value is the ratio of individuals diagnosed with the disorder in y-axis who also are diagnosed with the disorder in x-axis, in the subset of 1.7 million siblings for whom education data was available in the Dutch population registry. NA: missing value due to low sample size. Data in **Supplementary Table 7**.

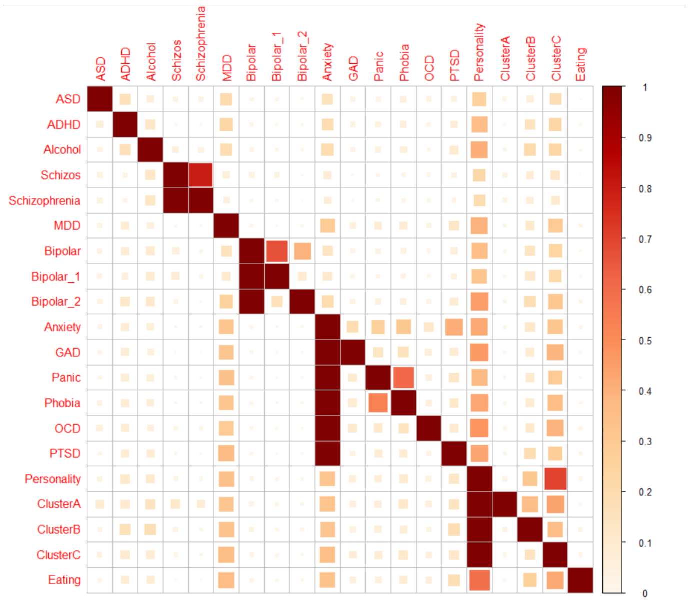

**B. Polychoric correlations**

Polychoric correlations between diagnoses in the subset of 1.7 million siblings for whom education data was available in the Dutch population registry. Anorexia nervosa was combined with bulimia nervosa into an eating disorder group due to the small number of individuals for either anorexia or bulimia diagnosis. NA: missing value due to low sample size. Data in **Supplementary Table 7**.

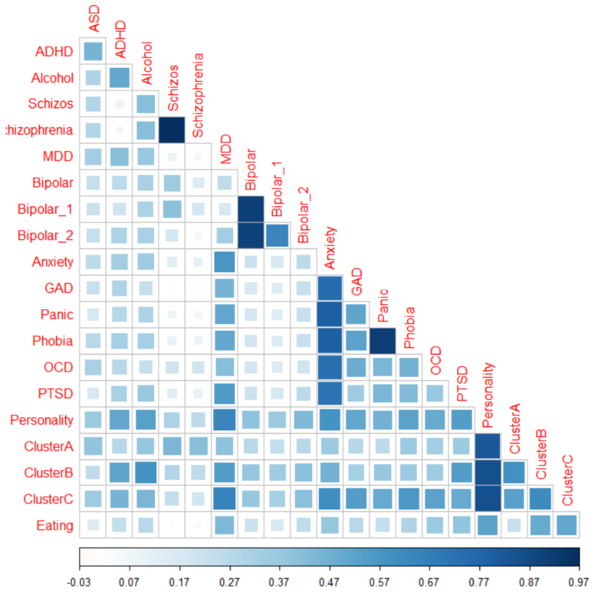

#### C. Genetic correlations

Genetic correlations between diagnoses estimated from summary statistics of the GWASs used in the MR analyses. Data in **Supplementary Table 8**.

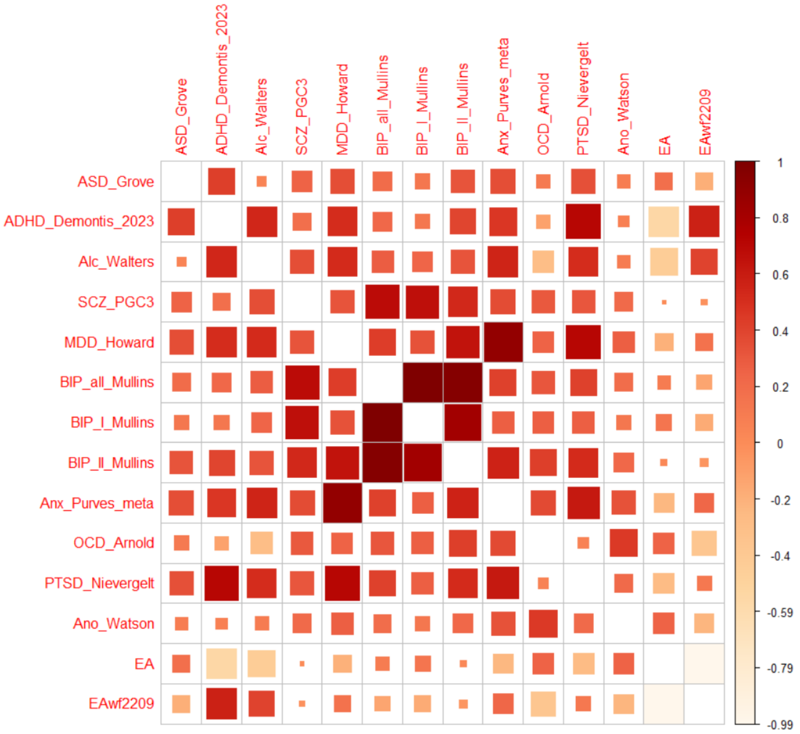

#### Supplementary Figure 3. Prevalence stratified by education and sex

Prevalence (expressed in percentage) of diagnoses in the sample of siblings for whom education data was available stratified by disorder (panels), education (x-axis) and sex (colour). Bars represent 95% confidence intervals. Note the scales of the y-axis are adapted depending on the diagnosis. In this figure, anorexia nervosa was combined with bulimia nervosa into an eating disorder group due to the low sample size of diagnosed individuals for some EA/sex strata. Data in **Supplementary Table 9**.

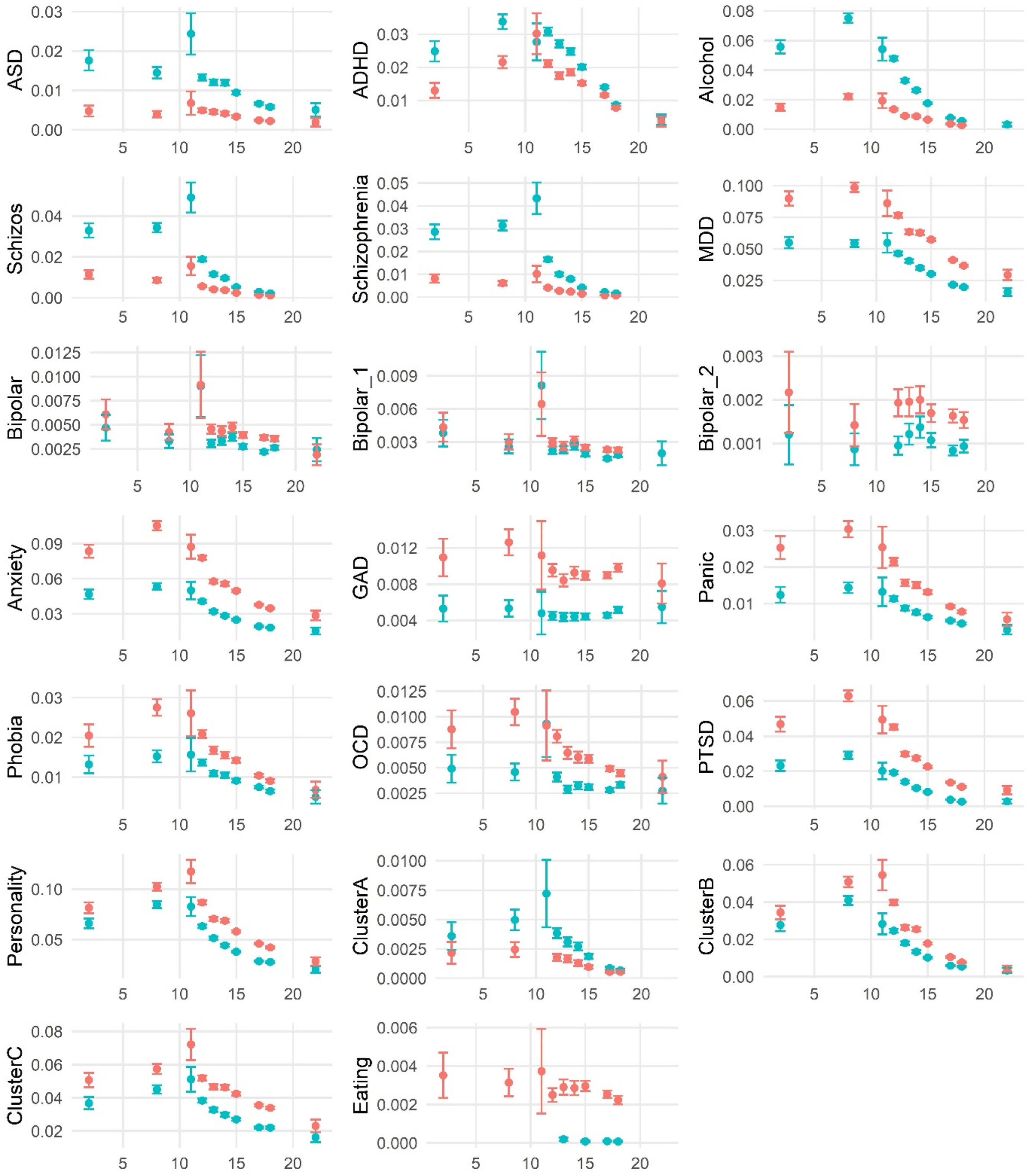

#### Supplementary Figure 4. Within-sibship analyses, excluding 2 and 11 years of education

Odds ratios per year of education as estimated with logistic regression (black) and within-sibship models (purple) in the Dutch population register, with all of sample (4) (left), when excluding siblings with 11 years of education (middle), and when excluding siblings with 2 years of education (right). Bars: 95% CIs. Data in **Supplementary Tables 10-13 & 16-17**.

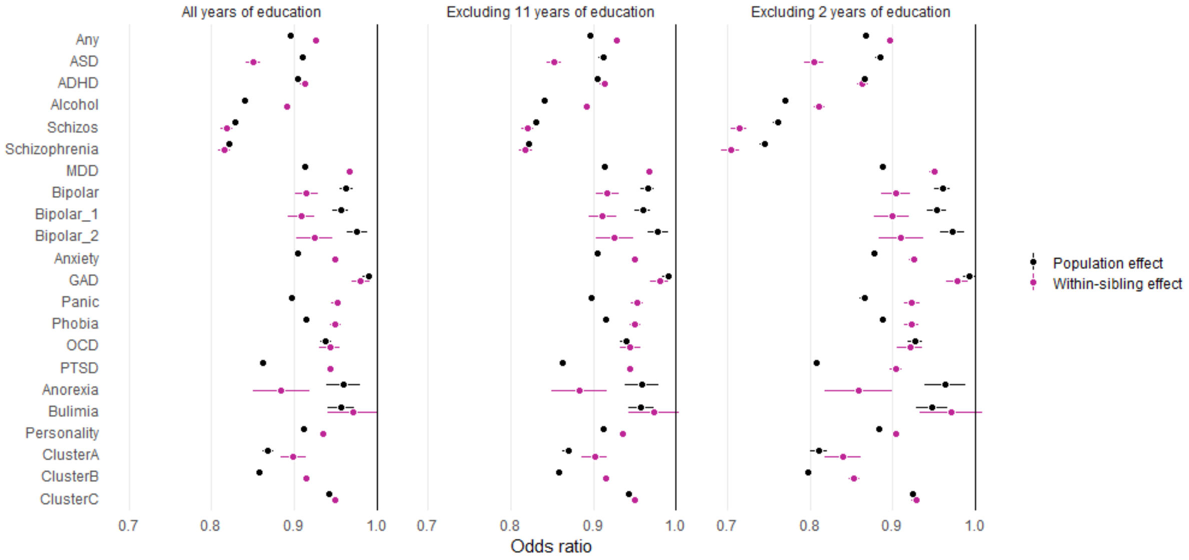

#### Supplementary Figure 5. Within-sibship analyses, sex-stratified

Odds ratios per year of education as estimated with logistic regression (black) and within-sibship models (purple) in the Dutch population register, in men-only sibships (left) and women-only sibships (right). Bars: 95% CIs. Data in **Supplementary Tables 14-15**.

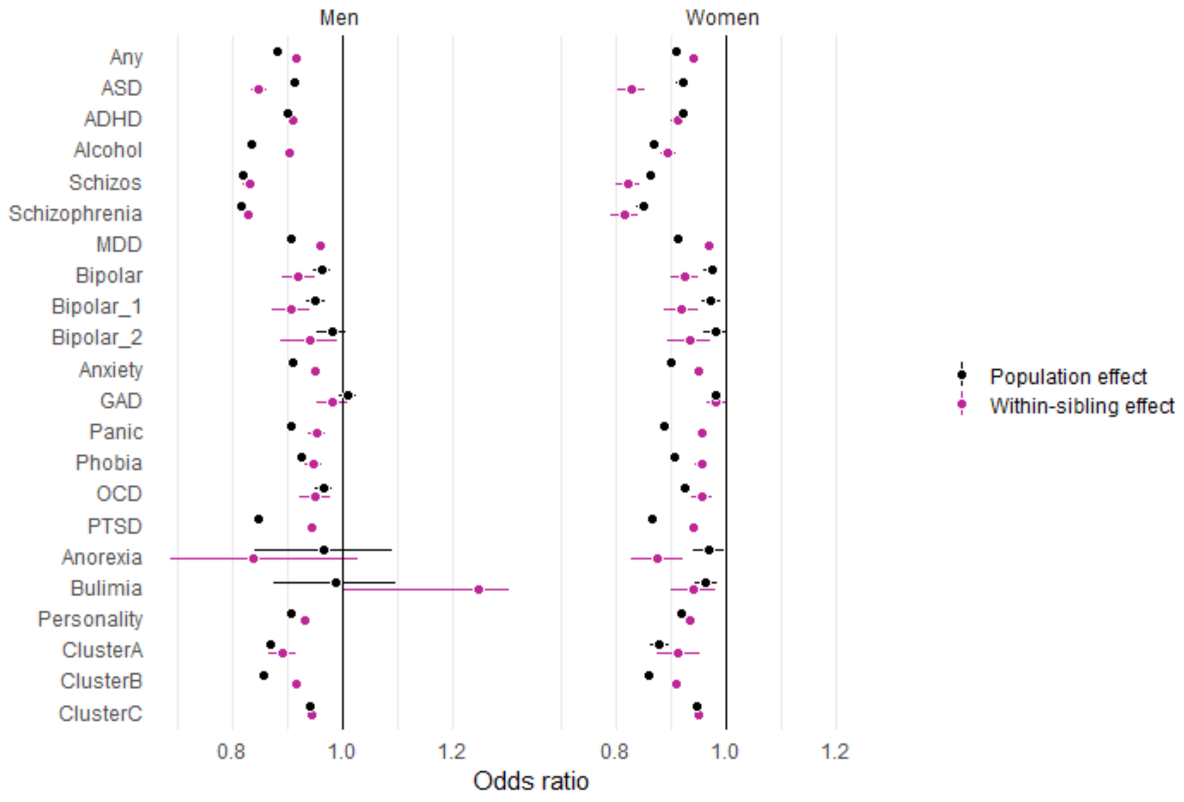

#### Supplementary Figure 6. Histogram of mental health expenditures per individual

X-axis is the average mental care expenditures spent between 2009-2018, in euros. Data in **Supplementary Table 18**.

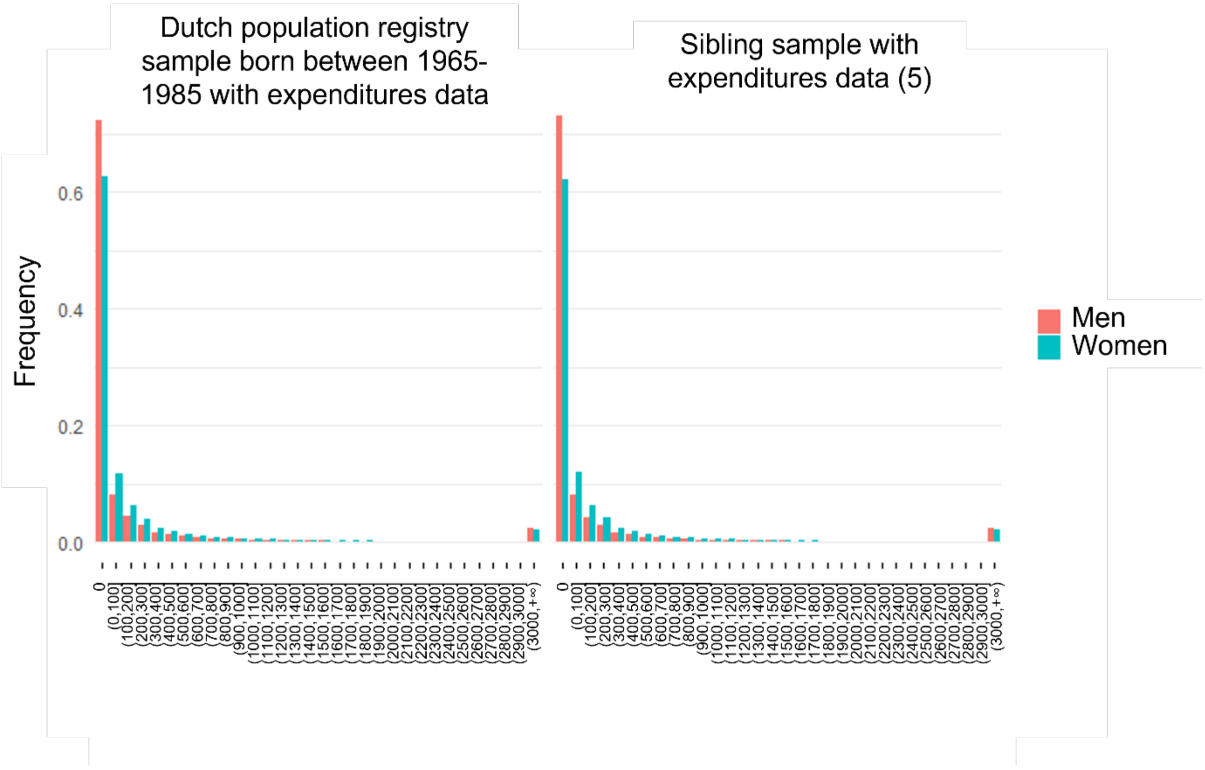

#### Supplementary Figure 7. Two-sample MR for all traits

**A. MR analyses of EA on diagnoses**

Odds ratios per year of education as estimated with the two-sample MR analyses of EA on diagnoses. Bars: 95% CIs. Data in **Supplementary Tables 22-23**.

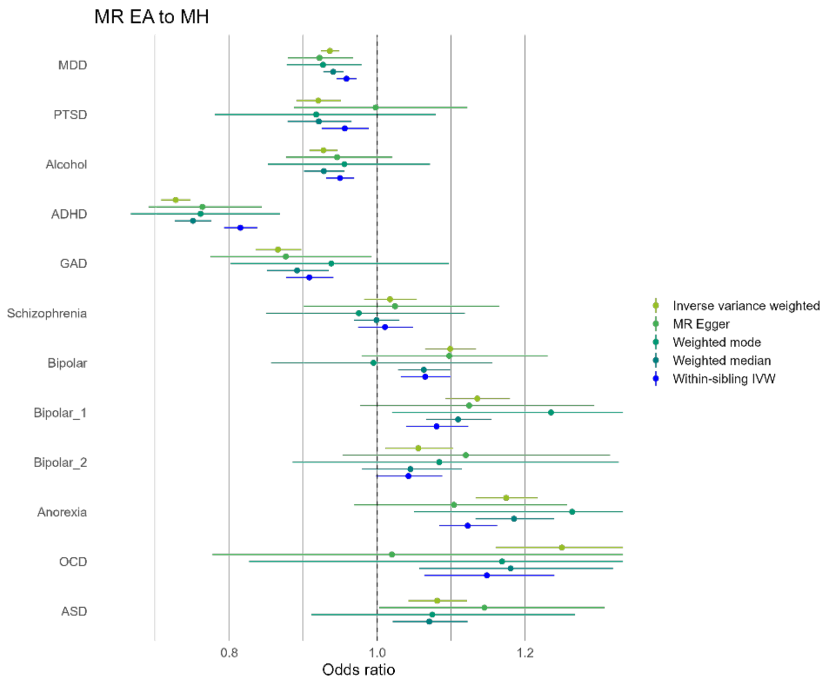

B. **MR analyses of EA on diagnoses**

Effect estimates of the two-sample MR analyses of EA on diagnoses. Bars: 95% CIs. Data in **Supplementary Tables 24-25**.

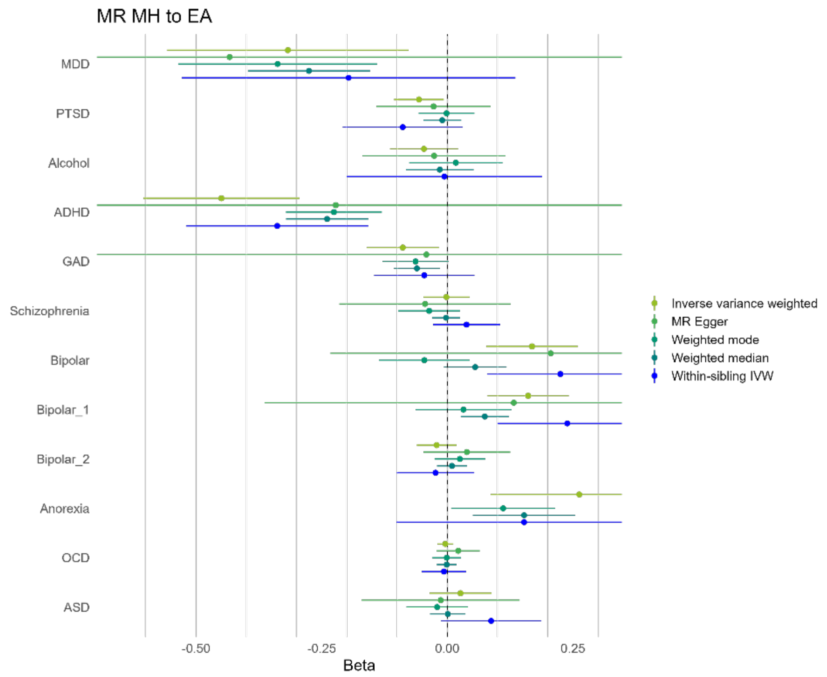

#### Supplementary Figure 8. Average number of years of education of patients, of healthy siblings of patients and unaffected sibships.

The red line is the average number of years of education of unaffected sibships (15.54 years of education), in families in which none of the siblings was diagnosed with a mental disorder between 2011 and 2016. Green dots are the mean education of affected siblings per diagnosis. Orange dots are the mean education of siblings of an affected sibling that are themselves were never diagnosed in the 2011-2016 timeframe. Bars: 95% CIs. Data in **Supplementary Table 26.**

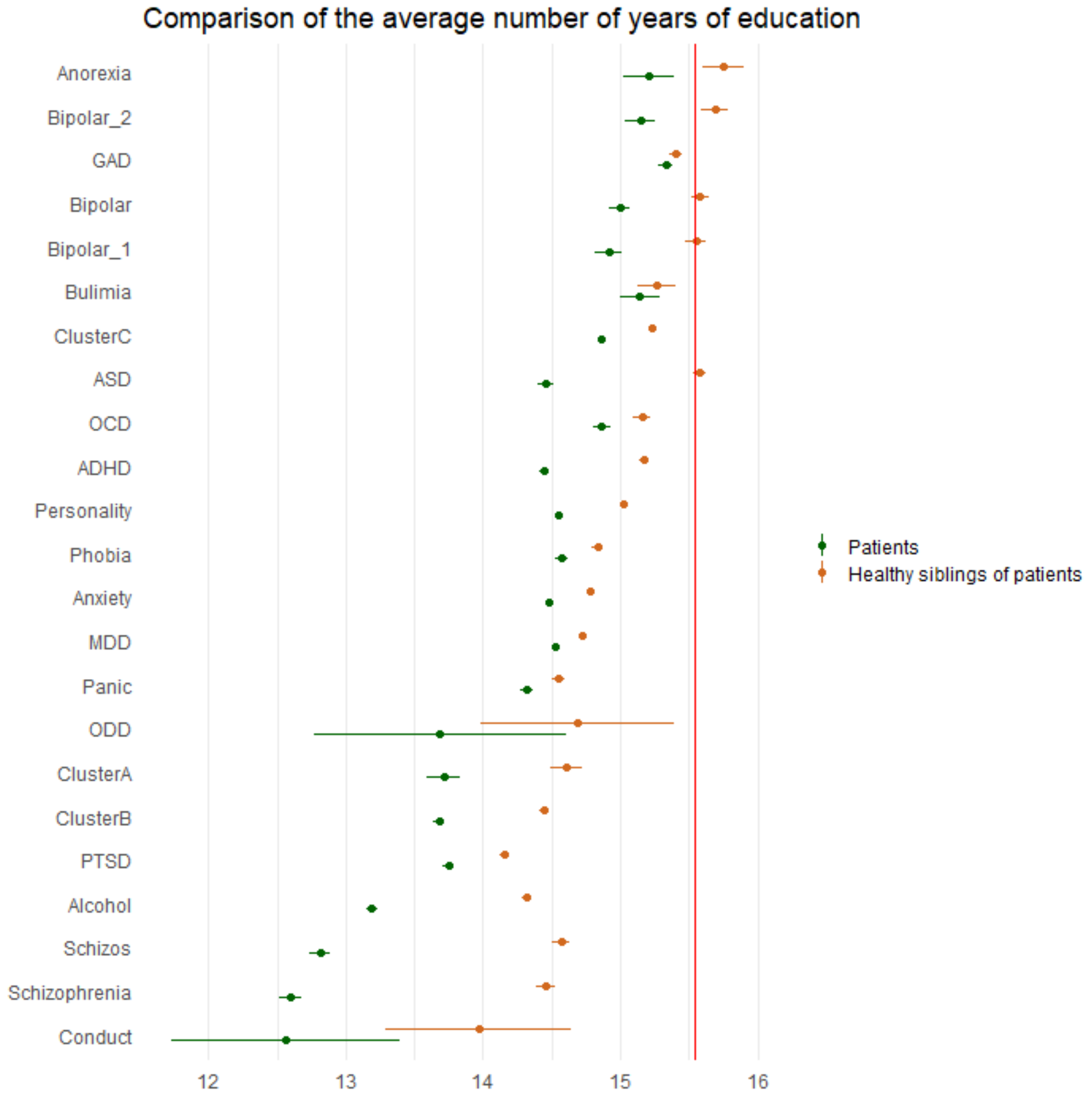
